# A New Algorithm for Incidental Pancreatic Cyst Detection

**DOI:** 10.1101/2024.09.18.24313888

**Authors:** M. Álvaro Berbís, Juan Moreno-Vedia, Félix Paulano-Godino, Ainhoa Viteri, Meritxell Riera-Marín, Daniel Cañadas-Gómez, Romina Trotta, Beatriz Forastero, Luis Luna, Javier García López, Antonio Luna, Júlia Rodríguez-Comas

**Affiliations:** Department of Radiology, HT Médica, San Juan de Dios Hospital, 14012 Córdoba, Spain; Scientific and Technical department, Sycai Technologies S.L., 08018 Barcelona, Spain; Department of Radiology, HT Médica, Clínica las Nieves, 23007, Jaén, Spain

**Keywords:** Pancreatic Cysts, Computerized Tomography, Magnetic Resonance Imaging, Incidental Findings, Artificial Intelligence, SwinT-Unet

## Abstract

**Objectives:** To develop an accurate, state-of-the-art algorithm for the incidental detection of pancreatic cystic lesions (PCLs) on computerized tomography (CT) and magnetic resonance imaging (MRI) scans.

**Methods:** A SwinT-Unet-based architecture was developed for the incidental detection of PCLs. The algorithm was trained and validated on a robust dataset of retrospective CT and MRI studies collected from HT Médica centers located in eight different cities using scanners fabricated by four different manufacturers.

**Results:** Our algorithm was able to detect 91.6% of the confirmed PCLs in the initial dataset with 91.6% sensitivity and 92.3% specificity, while 91.7% of the healthy controls were also correctly identified. Furthermore, our tool was remarkably capable of classifying these PCLs as mucinous or non-mucinous, determining their location within the pancreas with an accuracy of 88.5%, and identifying the presence of calcifications or scars within the PCLs with an accuracy of 96%.

**Conclusions:** By integrating radiological data and state-of-the-art artificial intelligence techniques, we have developed an efficient tool for the incidental identification and initial characterization of PCLs, which present a substantial prevalence within the global population. Our algorithm facilitates early diagnosis of pancreatic abnormalities, which could have a profound impact on patient management and prognosis, particularly in the case of PCLs with malignant potential.

## INTRODUCTION

Pancreatic cancer (PC) currently ranks as the twelfth most common type of cancer globally [1], and it is further expected to become the second leading cause of cancer-related mortality by 2030 [2]. Moreover, PC is one of the most lethal types of cancer, with a 5-year survival rate inferior to 10% [3]. The delayed onset of PC symptoms, often appearing when metastasis has already occurred, results in up to 85% of patients no longer being eligible for surgical resection, which has a profound negative impact on their prognosis [4,5].

Many research efforts are currently focused on the identification of biomarkers for early PC detection, aiming to improve patient outcomes. However, only one of these biomarkers, the serum carbohydrate antigen 19-9, has been approved by the US Food and Drug Administration (FDA), and only as a therapy response and disease relapse monitoring marker, as its predictive value is too low for population screening purposes [6]. Nonetheless, there is an opportunity for early PC diagnosis in patients presenting pancreatic cystic lesions (PCLs), as some of them are well-known precursors for this malignancy. The widespread adoption of advanced imaging techniques, particularly computed tomography (CT) and magnetic resonance imaging (MRI), has led to an increased identification of conditions unrelated to the initially suspected diagnosis. These incidental findings can indeed be fortunate discoveries when they entail the early detection of a potentially treatable malignancy. Unsuspected PCLs have a prevalence of 2.6% in CT scans [7] and of 13.5– 19.6% in MRI studies [8,9], and both show a strong correlation with advanced age. Unexpectedly, a recent study found a much higher prevalence (49.1%) of incidental PCLs in healthy individuals, which also increased with body mass index and age [10].

PCLs can have a non-neoplastic (pseudocysts) or a neoplastic nature. Among the latter, serous cystadenomas (SCA) are typically regarded as benign [11], while mucinous cysts are often associated with malignant potential. Intraductal papillary mucinous neoplasms (IPMN) and mucinous cystic neoplasms (MCN) are two well-known PC precursors. IPMNs can arise from the side branches, the main pancreatic duct, or a combination of both, with a notably higher prevalence of PC found in main duct IPMNs [12]. Thus, this distinction has a direct impact on patient management and prognosis. PC cases arising from PCLs have been proposed to follow a systematic model in which malignancy progression occurs over several years, thus offering an opportunity for early diagnosis [13,14]. However, differentiation between the different types of PCLs is challenging, and although the presence of some specific features can be indicative of malignancy [15,16], these signs are sometimes not enough to confidently distinguish between benign and malignant PC precursors. Nevertheless, early detection of PCLs is essential to improve patient outcomes and reduce the economic strain on healthcare systems, as it would allow more informed, enhanced decision-making regarding lesion management and monitoring, thus potentially preventing progression to PC.

Artificial intelligence (AI) tools are poised to play a crucial role in the early detection of PCLs in CT and MR images. These algorithms have the potential to accurately identify and define lesion boundaries and to extract essential qualitative and quantitative information from their features, thus improving diagnostic precision. The implementation of these tools would also help to streamline the workflow in radiology departments, providing valuable assistance to radiologists in the diagnostic process and reducing their workload. The capability of convolutional neural networks (CNNs) to learn the spatial hierarchies of features from input data in an automatic and adaptive manner makes them exceptionally useful for extracting information from medical images. This capability allows CNNs to learn complex patterns at different levels of abstraction, recognize patterns independently of their spatial location, and even identify new patterns not obvious to the human eye. Moreover, depending on the specific dataset they are trained on, CNNs can work with multiple types of imaging studies (ultrasound, CT, MRI, etc.) and perform a wide variety of tasks (detection, segmentation, classification, etc.). Applied to medical images, the AI model presented in this manuscript, based on the SwinT-Unet architecture, offers exceptional segmentation accuracy and remarkable generalization ability, allowing the discrimination of structures at the pixel level. The results showed an outstanding performance of our algorithm, achieving 91.6% sensitivity and 92.3% specificity in the incidental detection of PCLs, while also demonstrating a remarkable capacity to further characterize the lesion by classifying it as mucinous or non-mucinous, accurately determining its location within the pancreas, and identifying the presence of calcifications or scars within the PCLs. Our findings confirm a significant prevalence of PCLs within our study population, highlighting the crucial support a tool like ours might provide through the early diagnosis of pancreatic abnormalities and the significant impact this would have on patient management and prognosis, particularly in the case of PLCs with malignant potential.

## MATERIALS AND METHODS

### Patients

The total cohort included 43.3% women (**table 1**). This initial cohort was divided into two subgroups: the control group included 56.3% of the patients (37.9% women, **table 2**), while the second group, comprising patients who had been diagnosed with a PCL, included 43.7% of the study population (**table 3**). Both groups showed a similar age distribution, and women and men were equally represented in the PCL group, which was further characterized to analyze the performance of our algorithm.

**Table 1.** Demographic and manufacturer distribution of the total cases included in the study.

|  | **Total Cases** | ***p*** |
| --- | --- | --- |
| **Age (years)** | 66 (55-74) |  |
| 0-18 | 0 | **<0.001** |
| 19-40 | 3.7% |  |
| 41-60 | 32% |  |
| 61-80 | 60% |  |
| 81-100 | 4.3% |  |
| **Women** | 43.3% | **0.021** |
| **Manufacturer** |  | **<0.001** |
| Siemens | 47% |  |
| Philips | 26.3% |  |
| GE Medical systems | 15.7% |  |
| Canon Medical Systems | 11% |  |

**Table 2.**
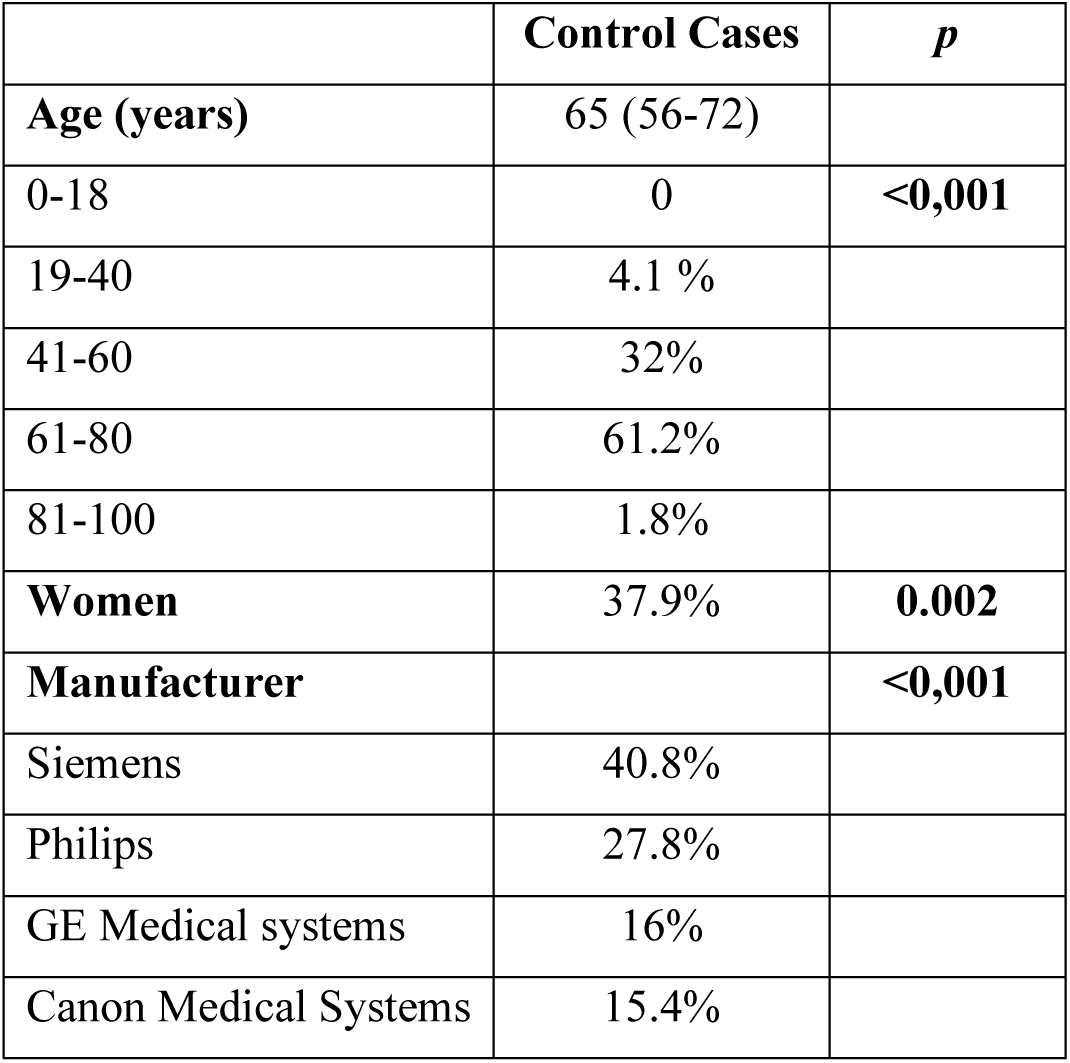
Demographic and manufacturer distribution of the control cases included in the study.

**Table 3.** Demographic and manufacturer distribution of the PCL cases included in the study.

|  | **PCL Cases** | ***p*** |
| --- | --- | --- |
| **Age (years)** | 68 (54-76) |  |
| 0-18 | 0 | **<0.001** |
| 19-40 | 3.1% |  |
| 41-60 | 32.1% |  |
| 61-80 | 57.3% |  |
| 81-100 | 7.6% |  |
| **Women** | 50.4% | 0.930 |
| **Manufacturer** |  | **<0.001** |
| Siemens | 55% |  |
| Philips | 24.4% |  |
| GE Medical systems | 15.3% |  |
| Canon Medical Systems | 5.3% |  |

PCL: Pancreatic Cystic Lesion

### Imaging

Image studies were acquired at different HT Médica medical centers located in the Spanish cities of Jaén, Córdoba, El Ejido, Huelva, Cádiz, Jerez de la Frontera, Algeciras, and Sevilla, from January 2018 to December 2021, using scanners developed by the following manufacturers (**tables 1–3**): Canon Medical Systems (Otawara, Japan), GE Healthcare (Chicago, IL, United States), Philips (Amsterdam, The Netherlands), and Siemens Healthineers (Erlangen, Germany). Demographic data of the patients, including age and gender, were collected from HT Médica’s radiological information system.

### Segmentation and feature extraction

A team of radiologists, each of them with more than five years of experience, manually delineated the pancreas and PCLs in all the images, slice by slice, using the 3D modeling tool available in the Philips IntelliSpace Portal (v12.1). The segmentations were subsequently exported in RTSTRUCT format. To ensure agreement between raters, collaborative segmentation, and consensus resolution were employed, minimizing discrepancies in the assessment process.

### Algorithm development

The proposed model (**fig 1**) is based on the SwinT-Unet [17] architecture, a further development from the U-Net [18] that aims to predict objects on an image with pixel-wise accuracy. Our model includes an encoder block, to reduce input resolution, and a U-Net decoder with dual-scale information modules. The encoder is an attention-based model with blocks that capture information at multiple scales, similar to that proposed by Atek et al. [17]. The input image is processed through multiple Swin transformer blocks performing attention operations and feature transformation. These blocks are responsible for feature extraction at various scales, allowing the model to capture both the fine details and high-level abstract features. After information passes through the encoder, the architecture adopts a U-Net-like structure [18] to perform segmentation. The decoder takes the features extracted by the encoder and uses them to generate a segmentation mask of the same resolution as the original input. Additionally, the decoder incorporates dual-scale information modules to merge features from different resolution levels that allow the model to integrate detailed and contextual information at different scales, thus improving segmentation accuracy, especially in areas with fine details or small features. This ability to process features and key points at different levels is crucial for the network to learn the features that best characterize big organs, such as the liver, as well as small lesions with different shapes and textures, such as PCLs. The network was specifically trained to learn the position and shape of the liver, kidneys, and pancreas, as well as a wide range of benign, pre-malignant, and malignant lesions present in the training set corresponding to the aforementioned organs. The model proposed here further includes two more steps: (1) a pre-processing step, prior to the inference of the neural network, that normalizes the input image by applying a Soft-Tissue Normalization [19], and (2) a post-processing step after the inference the filters out potential detections of the network that have no anatomical meaning, such as lesions belonging to an organ which they are not in direct contact with or predictions present in parts of the study where the abdomen is not visible yet.

**Fig 1.**
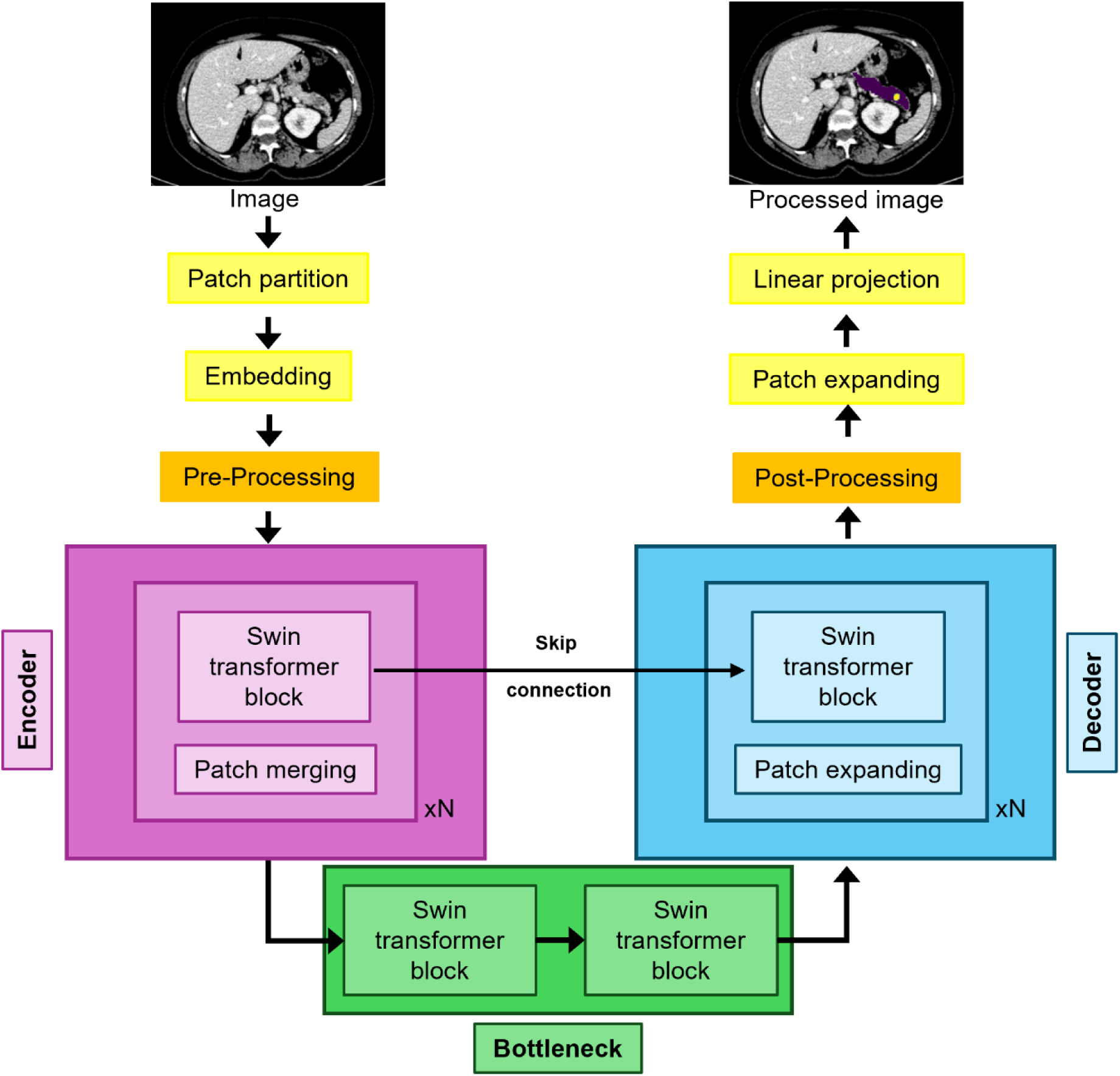
Schematic representation of the Swin-Unet architecture. The algorithm integrates a Swin transformer-based encoder and a symmetrical Swin transformer-based decoder, connected by two successive Swin transformer blocks (bottleneck).

### Statistical analysis

The Kolmogorov–Smirnov test was used to assess normality. Categorical variables are presented as total numbers and percentages, and continuous variables are presented as medians and interquartile ranges (IQRs) for non-normally distributed data. The chi-square (χ2) test was used to analyze group differences for categorical variables, while the Mann– Whitney U test was used for continuous variables. Results were considered statistically significant if *p*<0.05. Statistical analyses were conducted using the SPSS Statistics, version 29.0.0.0 (IBM, Armonk, NY, United States).

## RESULTS

### Patient’s characteristics

The median age of the patients included in the present study was 66 years old. The 61–80 age range was the most represented, consistent with the expected increase in CT and MRI scans performed as the population ages. Women accounted for 43.3% of the initially evaluated patients (**table 1**). The recruited cohort was divided into two groups: one group with confirmed PCLs, including 43.7% of the patients, and a control group of healthy individuals. Both groups showed a similar age distribution, and women and men were equally represented in the PCL group (**tables 2 and 3**).

Among these PCL lesions, 61.1% of them were identified as non-mucinous (serous cystic neoplasms (SCN), pseudocysts) and 38.9% were classified as mucinous (MCNs, IPMNs). Within the non-mucinous group, 93.8% of them were SCNs or pseudocysts; additionally, 3.8% of non-subclassified benign lesions and 2.5% of undetermined non-mucinous lesions were detected (**table 4**). The prevalence of both mucinous and non-mucinous lesions increased with age, with mucinous cases associated with a higher median age (*p*<0.001). Notably, a higher percentage of patients with mucinous lesions fell within the 61–80 age range, consistent with the expectation of a higher lesion incidence in older patients [10].

**Table 4.** Clinical and demographic characteristics of mucinous and non-mucinous PCLs.

|  | **Mucinous PCL** | **Non-mucinous PCL** | ***p*** |
| --- | --- | --- | --- |
| SCN | - | 70% |  |
| Pseudocyst | - | 23.8% |  |
| Benign non-subclassifiable | - | 3.8% |  |
| Undetermined | - | 2.5% |  |
| IPMN | 90.2% | - |  |
| MCN | 9.8% | - |  |
| **Age (years)** | **72 (64-79)** | **63 (51-74)** | **<0.001** |
| 0-18 | 0 | 0 |  |
| 19-40 | 0 | 5% |  |
| 41-60 | 19.6% | 40% |  |
| 61-80 | 68.6% | 50% |  |
| 81-100 | 11.8% | 5% |  |
| **Women** | **54.9%** | **47.5%** | **0.409** |

PCL: Pancreatic Cystic Lesion; SCN: Serous Cystic Neoplasms; IPMN: Intraductal Papillary Mucinous Neoplasms; MCN: Mucinous Cystic Neoplasms.

### Incidental finding of PCLs

Out of the initial dataset, 43.7% of the cases were confirmed as true positives for a PCL, based on radiological evidence. This prevalence is in alignment with the existing literature, which reports the presence of PCLs in up to 49.1% of the adult population [10]. Our algorithm successfully detected 91.6% of the lesions, while 8.4% were missed (false negatives). Furthermore, among the control population, the algorithm accurately identified a healthy pancreas in 91.7% of the cases but presented 7.7% of false positives.

Overall, our SwinT-Unet-based model showed 91.6% sensitivity and 92.3% specificity in the detection of PCLs. These results confirm the high precision and reliability of our AI-based approach in distinguishing between cystic lesions and non-cystic structures within the pancreas, underscoring the potential of AI technology to enhance the accuracy and efficiency of pancreatic lesion detection, thereby facilitating early diagnosis and treatment planning for patients with pancreatic abnormalities.

### Characterization of PCLs: mucinous vs. non-mucinous classification and location

While the majority of pancreatic cysts carry a low risk of malignancy, some are recognized as premalignant lesions capable of progressing into mucin-producing adenocarcinoma. Consequently, the identification of these cysts often triggers heightened anxiety in the patient and prompts additional medical investigations to assess the potential for malignancy [20,21]. While the primary objective in this study was the incidental detection of PCLs, the classification of these cysts as mucinous (IPMNs, MCNs) or non-mucinous (SCNs, pseudocysts) [22] was also addressed, achieving an accuracy of 73.3% in the classification of PLCs as mucinous (**fig 2**).

**Fig 2.**
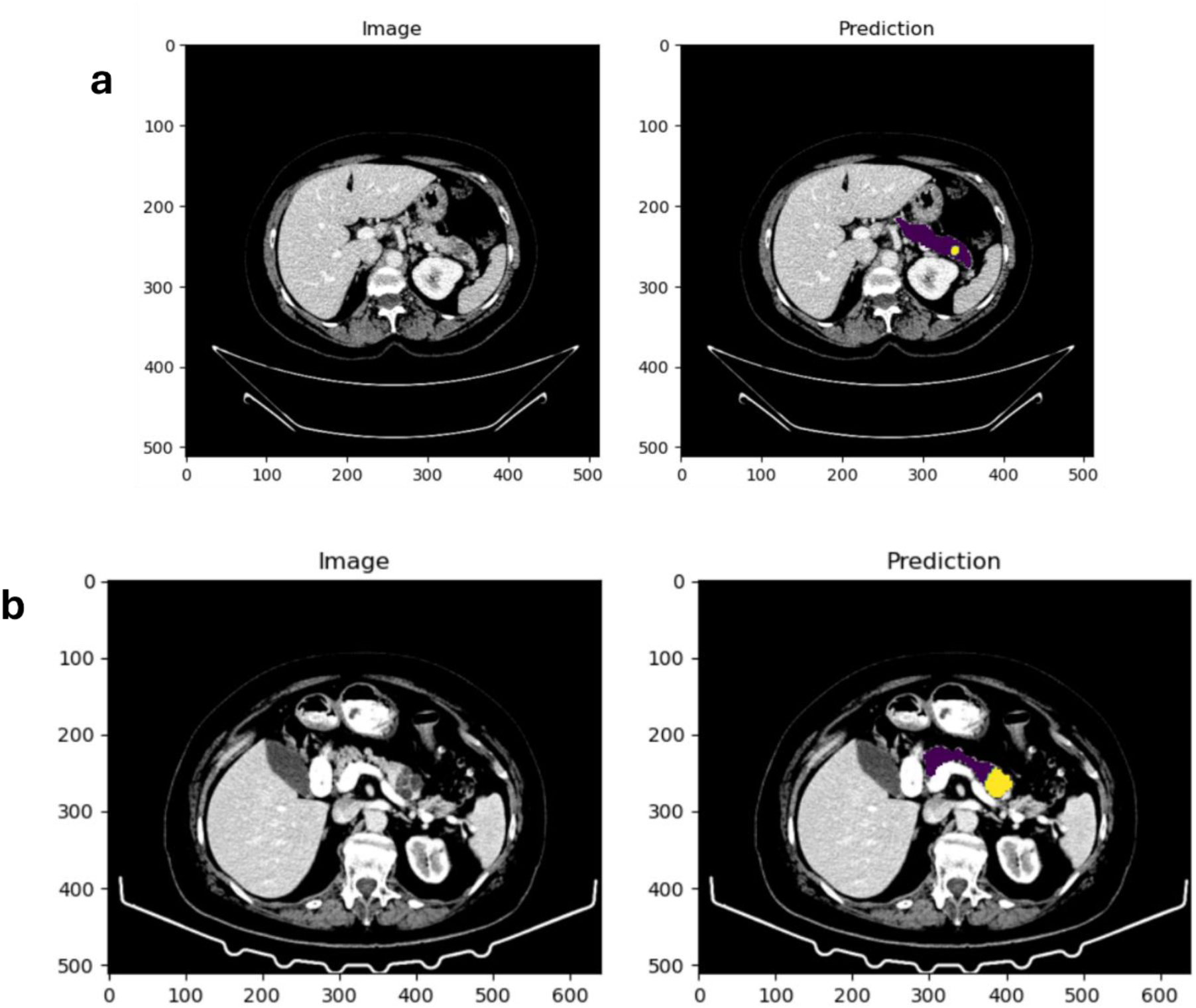
Original images and predictions made by our algorithm. (A) Mucinous PCL. (B) Non-mucinous PCL. Healthy pancreas is shown in purple, while the lesions appear in yellow.

Furthermore, we analyzed the spatial distribution of the PCLs within the pancreas, with the algorithm demonstrating a high accuracy (88.5%) in categorizing the cysts as head, body, or tail, according to their location. Specifically, the algorithm was capable of correctly determining the location of the lesion at the head or the uncinate process of the pancreas with an accuracy of 86%, at the body with an accuracy of 92.6%, and at the tail with an accuracy of 75.7%.

Additionally, we conducted a thorough examination for the presence of calcifications or scars within the cystic lesions, achieving an accuracy of 96% in their identification. This was crucial to assess the presence of potential signs of malignancy or chronic inflammation within the PCL. To achieve this, several image feature extraction algorithms were applied to study the high increments of Hounsfield Units (HU) on small sliding windows passed through patches of the image where the lesion was detected. A map of the increased direction of the HU within the sliding window is thus generated and, by calculating its maximum, the candidates to potential scars (understood as groups of pixels with a high HU, since they have a great bone component that is aligned towards a defined direction within the PCL) are extracted.

This comprehensive characterization of the PCL provides valuable insights into the diverse nature of the lesion, supporting clinicians in diagnostic decision-making and risk stratification for further management strategies.

## DISCUSSION

Considerable efforts have been employed to try to distinguish between the different types of PCLs, as this step is essential to correctly stratify the malignant potential of the lesion so the best patient management can be provided. Duh et al. developed an AG-Net model capable of identifying PCLs on CT scans with 93.1% sensitivity and 81.8% specificity, further classifying them into two groups (IPMN and MCN vs pseudocysts and SCAs) [23]. Vilas-Boas et al. designed a CNN for the automatic detection of mucinous PCLs. The algorithm, trained on images retrieved from EUS examination videos, achieved 98.5% accuracy, 98.3% sensitivity, and 98.9% specificity in the classification of PCLs as mucinous or non-mucinous [24]. Yang et al. developed a random forest (RF) model capable of differentiating between serous and mucinous pancreatic cystadenomas based on the analysis of radiomics texture features extracted from CT scans. Their algorithm achieved 0.83 accuracy, 0.85 sensitivity, and 0.83 specificity for a slice thickness of 5 mm [25]. Shen et al. compared the performance of a support vector machine (SVM) model, an RF algorithm, and an artificial neural network (ANN) in differentiating among SCAs, MCNs, and IPMNs using eight clinical factors and nine radiomics features extracted from CT scans [26]. The RF classifier offered the best results, achieving an accuracy of 79.59% and F1 scores of 0.7500 for the differentiation of IPMNs, 0.8182 for MCNs, and 0.8077 for SCAs. Gao et al. designed a CNN to identify pancreatic anomalies, including PCLs, on MRI images [27]. The authors employed 504 original pre-treatment MRI studies to train their model. As most of the patches within the images corresponded to carcinoma, they augmented the number of images for the other conditions with the help of a generative adversarial network (GAN), creating synthetic images based on real ones up to a total of 35735 patches for the training dataset. The CNN trained on this augmented dataset offered its best results when a synthetic to real images ratio of 40:1 was used, achieving an AUC of 0.9147 for the identification of carcinomas, 0.8486 for benign ductal diseases, 0.9126 for benign cystic diseases, 0.7189 for inflammatory diseases, 0.9301 for pancreatic neuroendocrine tumors, and 0.8880 for solid pseudopapillary tumors.

The number of studies exploring the incidental finding of PCLs via AI approaches is much reduced. Kooragayala et al. employed a publicly available natural language processing (NLP) software to identify incidental findings on CT scan reports [28]. The authors created a list of specific terms (including IPMN, pancreatic cyst, and pancreatic ductal dilation) to identify pancreatic findings that were used as parameters to train the algorithm on a subset of 28 patients who had undergone pancreatic resection for known pancreatic lesions. This algorithm achieved an accuracy of 0.987 on a validation set of 400 CT scan reports. Their optimized model was subsequently applied to 18769 CT studies from patients admitted at their institution for trauma and findings of interest were identified in 232 of them, including potential IMPNs (48 patients), pancreatic cysts (36 patients), concerning masses (30 patients), traumatic findings (44 patients), pancreatitis (41 patients), and ductal abnormalities (19 patients).

By utilizing CT and MRI scans originally intended for the identification or evaluation of conditions not related to pancreatic abnormalities, the algorithm we present in this study offers a novel approach to the detection of PCLs. Our tool confirmed the incidental presence of PCLs in 43.7% of the study population, in accordance with previously reported data [10]. Out of all confirmed cases, the SwinT-Unet-based algorithm correctly identified 91.6% of them with a remarkable 91.6% sensitivity and 92.3% specificity, improving the results reported by Duh et al., which were obtained with an algorithm specifically designed for the classification of PCLs, not for their incidental detection. Furthermore, our algorithm is capable of providing a comprehensive characterization of the PCL through its classification as mucinous or non-mucinous with 73.3% accuracy. Although this result did not improve those previously reported [24–27], the tool presented here can further characterize the PCL by categorizing the lesion according to its location in the head, body, or tail of the pancreas with 88.5% accuracy, as well as identifying the presence of calcifications or scars within the PCL with 96% accuracy. Taken together, all this information provides a comprehensive characterization of the lesion that, without any doubt, will be very valuable to clinicians for the planning of tailored patient management strategies.

The results presented in this study were achieved using a robust dataset consisting of CT and MRI studies obtained from eight different medical centers using scanners fabricated by four different manufacturers. However, they are limited by the somewhat reduced number of imaging studies included.

## CONCLUSIONS

Our study presents comprehensive findings regarding the incidental detection and characterization of PCLs through the integration of radiological data and advanced AI techniques. The results reveal a substantial prevalence of PCLs within the studied population, highlighting the importance of thorough radiological evaluations in clinical practice.

Our AI model, based on the SwinT-Unet architecture, showed a remarkable performance in detecting PCLs with a sensitivity of 91.6% and a specificity of 92.3%. Furthermore, our study contributes to the in-depth characterization of PCLs by distinguishing between mucinous and non-mucinous types, aiding in risk stratification and clinical decision-making, and the identification of calcifications or scars within the cystic lesions, an aspect crucial for the assessment of potential signs of malignancy or chronic inflammation within the PCL. These outcomes not only emphasize the clinical utility of our approach but also highlight the potential of AI-driven approaches in enhancing the accuracy and efficiency of pancreatic lesion detection, which will undoubtedly have a profound impact on patient management and prognosis, particularly in the case of PCLs with malignant potential.

## Data Availability

The data used in this study are not openly available due to commercial and proprietary constraints. Access to the data may be granted by the corresponding author upon reasonable request and subject to confidentiality agreements.

## DECLARATIONS

### Ethics approval

This research was performed in accordance with the ethical standards in the 1964 Declaration of Helsinki. The protocol followed in this study was approved by the Institutional Review Board of HT Médica.

### Consent

Written informed consent was obtained from the patients at the time of the performance of the imaging study.

### Authors’ contributions

Study conception and design were performed by M. Álvaro Berbís and Júlia Rodríguez-Comas. Material preparation, data collection, and analysis were performed by M. Álvaro Berbís, Juan Moreno-Vedia, Félix Paulano-Godino, Ainhoa Viteri, Meritxell Riera-Marín, Daniel Cañadas-Gómez, Romina Trotta, Beatriz Forastero, Luis Luna, and Javier García López. Supervision was performed by M. Álvaro Berbís and Júlia Rodríguez-Comas. The first draft of the manuscript was written by M. Álvaro Berbís, Javier García López and Júlia Rodríguez-Comas. Manuscript review and editing was performed by Antonio Luna. All authors commented on previous versions of the manuscript. All authors read and approved the final manuscript.

